# Virtual Reality as a Tool for Empathy Training in Healthcare Education: A Protocol for a Systematic Review of Qualitative Studies

**DOI:** 10.1101/2025.06.20.25329987

**Authors:** Ravi Shankar, Fiona Devi, Xu Qian

**Author notes:** **Corresponding Author:** Dr Ravi Shankar; Research and Innovation, Medical Affairs, Alexandra Hospital, Singapore, Email correspondence.

## Abstract

**Background:** Virtual reality (VR) technology has emerged as a promising tool for experiential learning in healthcare education, with particular interest in its potential to foster empathy by allowing learners to embody the patient perspective. Empathy is crucial for therapeutic alliance, patient satisfaction, and health outcomes, yet research consistently documents its decline as students progress through medical training. The COVID-19 pandemic has accelerated the adoption of immersive technologies and highlighted the importance of humanistic care. However, the feasibility, acceptability, and impact of VR empathy interventions are not yet well established.

**Objectives:** This systematic review aims to comprehensively synthesize qualitative evidence on the use of VR for empathy training in healthcare education. The review seeks to understand the feasibility, acceptability, and perceived impact of VR empathy interventions from the first-hand perspectives of learners, educators, and VR developers. Specifically, it will explore how VR empathy interventions are designed and implemented, examine learner experiences and engagement factors, identify perceived impacts on empathy and related outcomes, investigate facilitators and barriers to curriculum integration, and synthesize stakeholder-identified implementation considerations and best practices.

**Methods and Analysis:** A comprehensive search will be conducted across nine electronic databases (PubMed, Web of Science, Embase, CINAHL, MEDLINE, The Cochrane Library, PsycINFO, ERIC, Scopus) from inception to June 2025, supplemented by gray literature sources. Empirical qualitative studies focusing on VR interventions to foster empathy among healthcare students and professionals will be included using the SPIDER framework for eligibility criteria. Two reviewers will independently screen studies, extract data using a standardized form, and appraise methodological quality using the Critical Appraisal Skills Programme (CASP) qualitative checklist. Thematic synthesis will be employed to analyze and interpret findings, moving beyond simple aggregation to generate new insights and hypotheses. The review will be conducted according to PRISMA and ENTREQ guidelines, with the protocol registered in PROSPERO (CRD42025637500).

**Discussion:** Understanding how to optimize VR empathy training in healthcare education is timely and significant given the persistent challenges in maintaining empathy throughout medical training and the accelerated adoption of digital technologies. This review will provide actionable insights to ensure the thoughtful development and implementation of VR as an empathy-fostering tool. By synthesizing granular insights into participant experiences across diverse educational contexts, the findings can inform evidence-based instructional design, professional development for educators, and institutional support for VR initiatives. The review will also identify current gaps and challenges to guide a strategic research agenda for maximizing the impact of this emerging educational technology in cultivating a healthcare workforce equipped with both technical skills and human capacities needed for person-centered care.

## Introduction

Empathy, the ability to sense, understand, and share the feelings of another, is a cornerstone of compassionate, patient-centered care. In the clinical context, empathy involves both cognitive and affective processes - intellectually grasping the patient’s situation, resonating with their emotions, and communicating that understanding back to the patient [1]. Empathic care has been associated with numerous benefits, including improved patient satisfaction, treatment adherence, clinical outcomes, and lower provider burnout [2-5].

However, research has consistently documented a troubling decline in empathy as students progress through medical training [6,7]. The demands and stressors of medical education, including time pressure, fatigue, and exposure to suffering, can erode students’ empathic capacities [8]. The biomedical focus of curricula and the technological intensity of modern healthcare may also divert attention away from humanistic aspects of care [9]. Counteracting this empathy decline has thus become a key priority for medical educators and institutions.

While many educational strategies have been used to teach empathy, from literature and narrative medicine to patient perspective-taking exercises, the continuing empathy deficit suggests a need for more immersive, experiential approaches [10,11]. Just as flight simulators revolutionized pilot training by providing realistic, embodied practice, virtual reality (VR) technology may hold untapped potential as a tool for empathy education [12].

Virtual reality (VR) refers to interactive, computer-generated environments that dynamically respond to a user’s movements, creating a powerful sense of presence or the feeling of “being there” within the simulated world [13]. What sets VR apart from other forms of multimedia are three defining features. First, immersion, wherein sensory input from the virtual environment replaces real-world stimuli, transporting users to an entirely different setting. Second, interaction, which allows users to engage with and manipulate virtual objects through naturalistic movements, thereby enhancing engagement and realism. Third, imagination, as VR environments are not limited by the physical constraints of the real world, enabling users to experience scenarios that would otherwise be impossible or impractical in real life [14].

In medical and health professions education, VR has been increasingly adopted for clinical skills training, such as practicing surgical techniques, conducting physical exams, or responding to emergency scenarios [15,16]. By providing standardized, low-stakes practice opportunities, VR can supplement traditional training and accelerate skill development. Learners can receive automated feedback, track their progress, and learn at their own pace [17]. VR also overcomes logistical barriers of physical simulation, such as the cost of equipment and facilities [18].

Beyond procedural training, however, VR affords unique potential for what Dyer and colleagues term “socioemotional education” - cultivating empathy, compassion, and interpersonal skills [19]. The immersive quality of VR lends itself to perspective-taking; learners can virtually step into the shoes of patients to viscerally experience their thoughts, feelings, and challenges [20]. For instance, a medical student could see through the eyes of an elderly patient struggling to navigate the hospital, or a nursing trainee could embody a patient receiving a frightening diagnosis. Such virtual experiences may evoke a level of emotional engagement and personal insight difficult to achieve through passive learning methods [21].

A growing number of qualitative studies have begun to explore learner and educator experiences with VR empathy interventions across healthcare disciplines. For example, Kyaw et al. interviewed medical and nursing students who role-played as patients with Alzheimer’s disease in a VR simulation, eliciting themes of heightened empathy and appreciation for the patient’s perspective [22]. Lin et al. analyzed reflective essays from dental students who experienced the journey of a pediatric patient with cleft lip/palate, revealing increased understanding of psychosocial impacts and motivation to provide patient-centered care [23]. Ma et al. conducted focus groups with mental health providers who used VR to simulate auditory hallucinations in schizophrenia, uncovering insights into VR’s capacity to reduce stigma and sharpen empathic listening skills [24].

Despite the burgeoning interest in VR empathy training, the field remains nascent. Most studies to date have been small-scale pilots or single-site initiatives. The comparative effectiveness of VR versus other empathy education approaches is unclear. Technical and logistical barriers to implementing VR at scale, as well as potential unintended consequences such as distress or reinforcement of stereotypes, remain underexplored. A systematic synthesis of the complex, context-dependent experiences and perspectives of VR users is needed to guide thoughtful instructional design and deployment.

This systematic review aims to comprehensively synthesize qualitative evidence on the use of virtual reality (VR) for empathy training in healthcare education. The overarching objective is to understand the feasibility, acceptability, and perceived impact of VR empathy interventions from the first-hand perspectives of learners, educators, and VR developers. Specifically, the review seeks to explore how VR empathy interventions are designed and implemented in healthcare education settings, including their key learning objectives, instructional strategies, and technological features. It will examine how healthcare students and professionals subjectively experience and engage with VR empathy simulations, as well as the factors that promote or hinder immersion and perspective-taking. The review also aims to identify the perceived impacts of these interventions on empathy, attitudes, knowledge, and behaviors, and how they compare to traditional educational approaches. Furthermore, it will investigate the facilitators and barriers to integrating VR empathy training into healthcare curricula from both learner and educator perspectives. Finally, the review will synthesize stakeholder-identified implementation considerations, best practices, and research priorities for advancing the use of VR in empathy training.

By synthesizing granular insights into participant experiences across diverse educational contexts, this review aims to provide an in-depth understanding of how VR empathy interventions work, for whom, and under what circumstances. The findings can inform evidence-based instructional design, professional development for educators, and institutional support for VR initiatives. Uncovering current gaps and challenges can also guide a strategic research agenda to maximize the impact of this emerging educational technology.

## Methods

This systematic review will be conducted in accordance with the Preferred Reporting Items for Systematic Reviews and Meta-Analyses (PRISMA) statement [25] and the Enhancing Transparency in Reporting the Synthesis of Qualitative Research (ENTREQ) framework [26]. The protocol has been registered in the International Prospective Register of Systematic Reviews (PROSPERO; registration number: CRD42025637500).

### Eligibility Criteria

The SPIDER (Sample, Phenomenon of Interest, Design, Evaluation, Research type) framework, developed specifically for qualitative evidence syntheses, will guide the eligibility criteria for this review [27]. The Sample will include healthcare students—such as those in medicine, nursing, pharmacy, dentistry, and allied health—at any level of training (undergraduate, graduate, or continuing education), as well as healthcare professionals in clinical practice. Studies involving educators or VR developers will also be included if their perspectives pertain to VR empathy training for healthcare learners. The Phenomenon of Interest focuses on the use of immersive VR simulations aimed at fostering empathy, compassion, and understanding of the patient perspective. Interventions that address other interpersonal skills such as communication or cultural competence will be eligible if empathy is an explicit component. However, studies involving non-immersive virtual experiences, such as video-based or online modules, will be excluded.

Regarding Design, the review will include empirical qualitative studies employing methodologies such as ethnography, phenomenology, grounded theory, and qualitative description. Qualitative components of mixed methods studies will also be included if relevant data can be extracted. In contrast, quantitative studies, opinion pieces, and theoretical papers will be excluded. For Evaluation, eligible studies must explore participants’ first-hand experiences, perceptions, attitudes, or reflections related to VR empathy interventions, using data collection methods such as interviews, focus groups, observations, reflective writing, or open-ended survey responses.

Under Research type, the review will include primary qualitative research studies published in peer-reviewed journals or as grey literature, such as dissertations and conference abstracts. Systematic reviews, scoping reviews, and meta-analyses will be excluded, although their reference lists will be screened for potentially eligible primary studies. No restrictions will be placed on the language, date or geographic location of the studies.

### Search Strategy

A comprehensive search will be undertaken across multiple electronic databases to identify relevant studies from their inception to June 2025. The databases to be searched include PubMed, Web of Science, Embase, CINAHL, MEDLINE, The Cochrane Library, PsycINFO, ERIC, and Scopus. This broad search strategy is designed to capture a diverse range of interdisciplinary literature spanning healthcare, psychology, education, and technology, ensuring a thorough and inclusive evidence base for the review.

The search strategy will be developed in consultation with a health sciences librarian and include combinations of keywords and controlled vocabulary terms (e.g., MeSH, Emtree) relating to four key concepts: (a) virtual reality, (b) empathy/compassion, (c) healthcare students/professionals, and (d) qualitative research. A preliminary search string is:

(((“virtual reality” OR “computer simulation” OR “immersive virtual*” OR “head mounted display*” OR “haptic*” OR “augment* reality” OR “mixed reality”)) AND (“empath*” OR “compassion*” OR “perspective taking” OR “theory of mind” OR “patient understanding”)) AND ((“medical student*” OR “nurs* student*” OR “pharmacy student*” OR “allied health student*” OR “healthcare profession*” OR “healthcare provider*” OR “medical education” OR “health professions education”)) AND ((“qualitative” OR “interview*” OR “focus group*” OR “ethnograph*” OR “phenomenolog* OR “grounded theory” OR “case stud*” OR “observ*” OR “experience*” OR “perception*” OR “attitude*” OR “view*” OR “reflection*” OR “perspective*”))

Searches will be filtered for English language, but no date limits will be applied. For databases that do not provide robust subject headings, keywords will be searched in all fields (e.g., title, abstract, author-supplied keywords, main subject headings).

To capture grey literature, we will search ProQuest Dissertations and Theses, conference proceedings (e.g., Medicine Meets Virtual Reality, IEEE Virtual Reality Conference), and clinical trial registries (ClinicalTrials.gov, WHO International Clinical Trials Registry Platform). The first 25 pages of Google Scholar results will also be screened. Finally, we will hand-search the reference lists of included studies and relevant reviews for additional eligible studies.

### Screening and Selection Process

Search results will be collated and de-duplicated using Covidence, a web-based platform for systematic review management. Two reviewers will independently screen the titles and abstracts of all unique records against the eligibility criteria. Studies deemed potentially relevant by either reviewer will proceed to full-text review.

The full texts of selected studies will be independently assessed by two reviewers. Reasons for exclusion at this stage will be documented. Disagreements between reviewers at any stage will be resolved through discussion or adjudication by a third reviewer. The study selection process will be reported using a PRISMA flow diagram.

### Data Extraction

A standardized data extraction form will be developed and piloted to systematically capture key information from each included study. Two reviewers will independently extract the data, and any discrepancies will be resolved through discussion to ensure accuracy and consistency. Data items to be extracted will include study characteristics such as the authors, year of publication, country, research questions or objectives, and the theoretical framework guiding the study. Participant characteristics will also be recorded, including sample size, age, gender, race or ethnicity, healthcare discipline, and level of training or professional practice. Information on the VR intervention will be collected, covering aspects such as learning objectives and content, technological features (e.g., display type, controllers, haptics), pedagogical design elements (such as pre-briefing, debriefing, and reflection prompts), as well as the duration, frequency, and setting of VR use. Study methods will be documented in terms of study design, data collection techniques, and data analysis approaches. Key findings will include the main themes, categories, or theories generated from the data, illustrative participant quotes, and the authors’ interpretations and conclusions. Additionally, we will extract any implications, recommendations, and limitations highlighted by the study authors. If any information is unclear or missing, study authors will be contacted for clarification.

### Quality Appraisal

Included studies will be critically appraised using the Critical Appraisal Skills Programme (CASP) Qualitative Research Checklist,[28] a widely used tool for assessing the methodological quality of qualitative studies. The checklist consists of 10 questions covering key domains such as appropriateness of the research design, recruitment strategy, data collection methods, researcher reflexivity, ethical issues, rigor of the analysis, and clarity and value of the findings.

Two reviewers will independently appraise each study, with disagreements resolved through discussion. Studies will not be excluded based on quality, as the goal is to comprehensively synthesize all available evidence. However, the appraisal results will inform data interpretation and confidence assessments, with greater weight given to findings from higher quality studies.

### Data Synthesis

Thematic synthesis, as described by Thomas and Harden,[29] will be used to analyze and interpret the data. This approach combines elements of meta-ethnography and grounded theory to generate new insights and hypotheses beyond a simple aggregation of findings.

The first stage involves line-by-line coding of the findings from each study, including both participant quotes and author interpretations. Codes will be inductively derived to capture the meaning and content of each text fragment. The second stage involves organizing the codes into descriptive themes that summarize the key concepts across studies. Finally, the descriptive themes will be further interpreted and abstracted to generate analytic themes that go beyond the primary studies to address the review questions.

The synthesis will be conducted collaboratively by two reviewers using NVivo qualitative data analysis software. The full author team will iteratively review and refine the codes and themes to ensure clarity, coherence, and fidelity to the original data. Throughout the process, we will maintain detailed memos to record observations, questions, and reflections.

We will explore similarities and differences in experiences and perceptions across different participant groups (e.g., students vs. professionals), healthcare disciplines, and types of VR interventions. Contextual factors that may influence the implementation and impact of VR empathy training, such as institutional support, technological infrastructure, and curricular integration, will also be examined.

## Discussion

Rationale for Review Empathy is a vital component of humanistic, patient-centered healthcare, yet there is a pressing need to bolster empathy training in health professions education. Current educational approaches, while valuable, have not sufficiently stemmed the tide of empathy erosion during medical training. VR technology, with its capacity for immersive, embodied perspective-taking, offers a promising avenue for fostering empathy. However, the nascent state of the evidence base, coupled with the rapid evolution of the technology, poses challenges for educators and institutions seeking to implement VR empathy interventions.

This review aims to provide a comprehensive, in-depth synthesis of learner, educator, and developer experiences with and perceptions of VR empathy training across healthcare disciplines. By going beyond surface-level effectiveness data to explore the nuanced, contextualized realities of VR use in educational settings, the review can generate actionable insights to guide the thoughtful design, implementation, and study of VR empathy interventions.

Strengths and Limitations A key strength of this review is the use of rigorous, transparent methods based on established guidelines for qualitative evidence synthesis. The SPIDER framework for eligibility criteria, developed specifically for qualitative reviews, will help ensure that the most relevant and appropriate studies are captured. The comprehensive search strategy, spanning nine databases and grey literature sources, will minimize the risk of missing important studies. The use of two independent reviewers for screening, data extraction, and quality appraisal will enhance the reliability and trustworthiness of the findings.

Additionally, the use of thematic synthesis for data analysis is well-suited to the review aim of generating new interpretive insights beyond a summary of primary study results. The collaborative, iterative nature of the synthesis process, with input from a multidisciplinary team, will help ensure a robust and inclusive interpretation.

However, there are also several limitations to note. First, the restriction to English language studies, while necessary for feasibility, may exclude valuable research from non-English speaking settings. Second, the qualitative focus of the review means that quantitative data on the effectiveness of VR empathy interventions will not be systematically assessed. However, a separate quantitative review is planned to address this question.

Third, the rapidly evolving nature of VR technology may make it challenging to compare interventions across studies conducted at different times. Careful attention will be paid to the specific technological features and affordances of each intervention during data extraction and synthesis. Finally, as with all qualitative research, the findings will be shaped by the subjective experiences and interpretations of both the primary study participants and the review team. Reflexive practices, such as keeping a decision audit trail and engaging in regular team debriefing, will help ensure transparency and trustworthiness.

Implications and Conclusion The COVID-19 pandemic has accelerated the adoption of digital technologies in healthcare and education, creating a window of opportunity for VR empathy training [30]. At the same time, the pandemic has underscored the vital importance of empathic, compassionate care in the face of suffering and uncertainty [31]. As we reimagine the future of health professions education, it is crucial that we critically examine the potential of emerging technologies like VR to foster humanistic values and skills.

This systematic review will provide a foundational evidence base to guide the design, implementation, and evaluation of VR empathy interventions in healthcare education. By synthesizing key stakeholder experiences across a range of settings, the review can help ensure that VR is leveraged in a way that is acceptable, feasible, and impactful for learners and educators. The findings can inform best practices for curricular integration, faculty development, and learner support, as well as identify priority areas for future research and development.

Ultimately, the goal is to harness the unique affordances of VR to cultivate a healthcare workforce equipped with both the technical skills and the human capacities needed to deliver high-quality, person-centered care. By immersing learners in the lived experiences of patients and fostering an ethic of care, VR empathy training holds promise as a transformative approach to nurturing the empathic imagination. As the field continues to evolve, this review will provide a roadmap for the journey ahead.

## Data Availability

This is a systematic review protocol. No primary data were collected for this study. The protocol has been registered in the International Prospective Register of Systematic Reviews (PROSPERO; registration number: CRD42025637500). Data extraction forms and analysis plans will be made available upon reasonable request to the authors once the review is completed.

## Appendix A Data Extraction Form

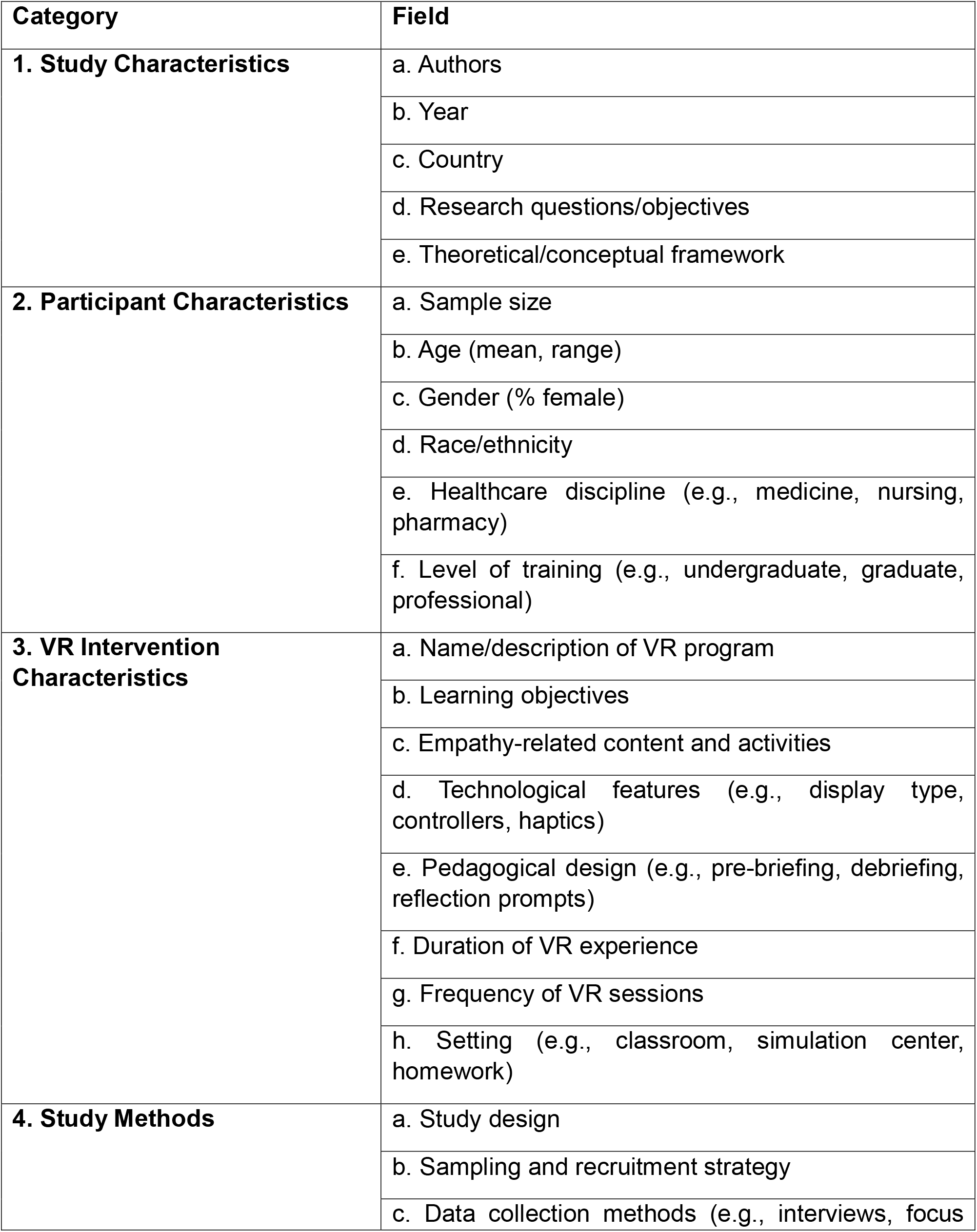
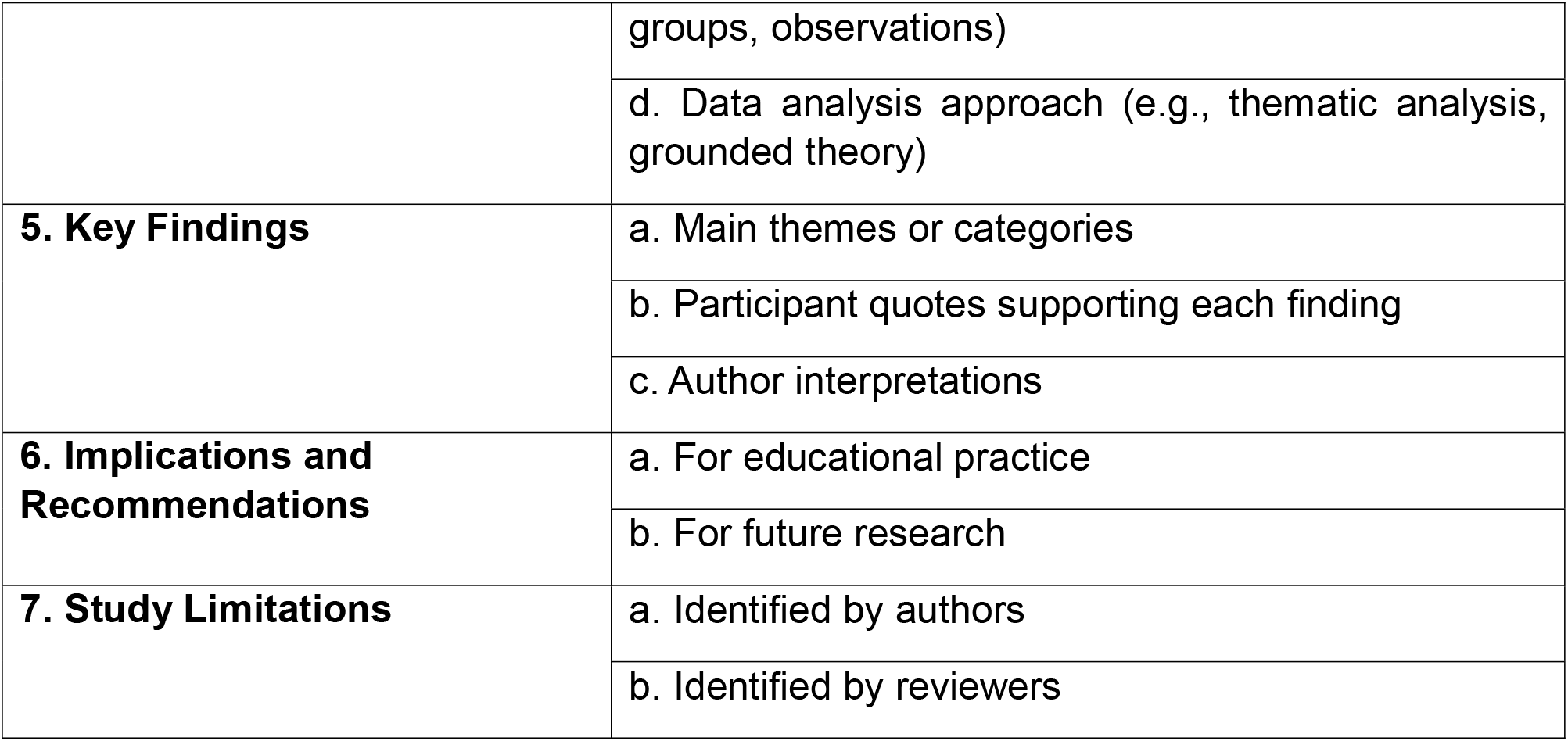

